# Personalized AI Prompt Generator and ChatGPT for Weight Loss: Randomized Controlled Trial in Adults with Overweight and Obesity

**DOI:** 10.1101/2025.09.07.25335255

**Authors:** Azwa Suraya Mohd Dan, Adam Linoby, Sazzli Shahlan Kasim, Sufyan Zaki, Razif Sazali, Yusandra Yusoff, Zulqarnain Nasir, Amrun Haziq Abidin

## Abstract

**Background:** The global prevalence of overweight and obesity continues escalating, driven by environmental factors and lifestyle behaviors leading to cardiovascular disease, diabetes, and cancer. Digital health interventions offer scalable solutions for weight management, yet personalized artificial intelligence applications remain underexplored. Large language model chatbots present opportunities for individualized behavioral interventions, but evidence comparing personalized versus manual prompt AI-driven approaches is limited.

**Objectives:** This randomized controlled trial evaluated the effectiveness of a personalized exercise and dietary prompt generator integrated with ChatGPT versus structured manual ChatGPT guidance for weight management.

**Methods:** Adults aged 18-65 years with BMI 27.5-34.9 kg/m^2^ (n=160) from Greater Kuala Lumpur were randomized to NExGEN personalized prompt system with ChatGPT (NEX; n=81) or structured manual ChatGPT control (CON; n=79). The NExGEN system generated individualized prompts based on 111 assessment variables including demographics, health status, and preferences. CON participants received structured manual ChatGPT prompts. Assessments occurred at baseline, 12 weeks, and 24 weeks. Primary outcome was body weight measured using bioelectrical impedance analysis. Secondary outcomes included body composition, dietary intake, physical activity, and cardiometabolic markers. Linear mixed models analyzed intervention effects.

**Results:** The NEX group achieved significantly greater weight loss than CON at 12 weeks (6.6 kg vs 3.0 kg, P<0.001) and 24 weeks (5.5 kg vs 1.7 kg, P<0.001). Fat mass decreased significantly in NEX participants (3.7 kg at 12 weeks, P<0.001) while preserving fat-free mass.

Energy density, fat intake, and physical activity improved significantly in the NEX group. Cardiometabolic variables showed minimal between-group differences.

**Conclusions:** The personalized AI-driven intervention produced superior weight loss and body composition improvements compared with structured manual prompt guidance. Limited cardiometabolic improvements reflected the metabolically healthy study population at baseline.

## 1. Introduction

Overweight and obesity have reached pandemic proportions worldwide, with recent estimates indicating that in 2021 over 1.0 billion men and 1.1 billion women globally were living with either overweight or obesity. Projections suggest that by 2050 more than half of the global adult population could be affected if current trends continue (Ng et al., 2025). In Southeast Asia and Malaysia, the escalating prevalence is particularly concerning. Malaysia now reportedly has one of the highest obesity rates in Southeast Asia, with over half of Malaysian adults being overweight or obese (Tee & Voon, 2024). These trends underscore an urgent public health challenge, as excess adiposity is a well-established risk factor for cardiometabolic conditions such as type 2 diabetes, hypertension, coronary heart disease, and metabolic syndrome (Stephenson et al., 2021). Epidemiological analyses attribute over 5 million deaths per year worldwide to high BMI, making obesity one of the leading preventable causes of death (Murray et al., 2020). The economic toll is likewise enormous, with recent analyses estimating that by 2035 the global economic impact will reach about $4.3 trillion annually (Mahase, 2023).

Conventional lifestyle interventions for weight management face well-documented challenges in both implementation and long-term adherence. Research indicates that a large majority of weight lost is often regained within 2-5 years after an intervention, due to both biological and behavioral factors (Varkevisser et al., 2019). Environmental and systemic factors further complicate individual efforts to manage weight through obesogenic environments characterized by abundant ultra-processed foods and sedentary living conditions (Swinburn et al., 2019). Traditional one-to-one counseling and in-person weight loss programs also have practical limitations in scalability and cost-effectiveness. Many primary care settings lack the capacity to provide the recommended intensive lifestyle therapy to every patient with obesity (Suojanen et al., 2020). These challenges have driven interest in greater personalization and new behavior change strategies to enhance traditional interventions.

In response to these limitations, there has been a surge of interest in digital health technologies to deliver lifestyle interventions for obesity at scale. Traditional face-to-face interventions often achieve 5-10% weight loss over 6-12 months, but require frequent meetings with healthcare professionals, making them labor-intensive and costly (Group, 2014). Digital health interventions offer a more scalable platform, with reviews and meta-analyses indicating they produce modest but significant weight loss, particularly when tailored (Beleigoli et al., 2019; Lau et al., 2020; Mamalaki et al., 2022; Protano et al., 2024). A growing subset incorporates artificial intelligence and machine learning to enhance personalization and user engagement. The advent of conversational AI chatbots for health coaching represents a notable development, with early studies showing promising outcomes (Stein & Brooks, 2017). Large Language Models like ChatGPT represent a cutting-edge frontier, demonstrating ability to engage in complex, context-aware dialogues for personalized health counseling (Ayers et al., 2023).

Current evidence suggests that AI-based interventions can facilitate behavior change, though effectiveness varies across studies. In a recent scoping review, the majority of AI chatbot interventions led to improvements in diet, physical activity, or weight outcomes (Chew, 2022). However, critical gaps remain regarding long-term durability and the degree of personalization needed for optimal effectiveness. Achieving effective personalization with AI may hinge on the quality of input data and prompt engineering, as large language model chatbots generate responses based on the prompts and information given to them (Teepapal, 2025; Wang et al., 2024).

Recent efforts have focused on developing personalized prompt generation systems that can tailor interventions to each individual’s profile in real time. The Nutrition and Exercise Planning Artificial Intelligence Prompt Generator (NExGEN) represents such an innovation, designed to collect comprehensive lifestyle assessment data and generate personalized prompts for AI-driven chatbot coaching. This approach addresses the critical gap between generic AI advice and truly individualized health coaching by systematically capturing user-specific variables to inform tailored recommendations. Studies are beginning to test these multi-component digital interventions, with preliminary results showing greater weight loss in groups receiving personalized AI coaching compared to standard digital programs (Zhou et al., 2018). The incorporation of dietary energy density concepts is particularly noteworthy, as AI can analyze food logs to identify opportunities for beneficial substitutions (Most et al., 2019). Recent randomized controlled trials have demonstrated the potential of personalized AI-guided interventions for weight management, with AI-driven approaches showing superior weight loss outcomes compared to standard care (Pokushalov et al., 2025). Despite these promising developments, no prior trial has combined a personalized prompt generator with AI-chatbot platform integrating both exercise and dietary strategies. The purpose of this randomized controlled clinical trial was to investigate the effectiveness of a personalized exercise and dietary prompt generator (i.e., NExGEN) integrated with ChatGPT compared to a structured manual ChatGPT control program for weight management in adults with overweight and obesity. This study hypothesized that the personalized program would result in statistically significant improvements with small to medium effect sizes, with these improvements being significantly more pronounced than in the control program.

## 2. Methods

### 2.1 Study Protocol

This study examined the comparative effectiveness of a personalized AI-driven prompt generator system (NExGEN) integrated with ChatGPT versus a structured manual ChatGPT control program for weight management in adults with overweight and obesity. A detailed description of the study intervention has been registered at the UMIN Trials Registry (registration number UMIN000053570). Additional information on the NExGEN system development and validation procedures can be found in the supplementary technical documentation.

### 2.2 Study Design

Study participants residing in Greater Kuala Lumpur (Klang Valley), Malaysia, who expressed interest through online and offline recruitment channels were enrolled in this randomized controlled clinical trial between July 2024 and December 2024. Participants attended in-person assessments where comprehensive anthropometric, body composition, dietary, physical activity, and cardiometabolic parameters were systematically collected by trained research personnel using standardized protocols. Clinical assessments were conducted at baseline (week 0), post-intervention (week 12), and follow-up (week 24) to evaluate the sustained effects of the interventions on primary and secondary outcome measures. During these clinical assessments, participants had the opportunity to provide qualitative feedback regarding their experience with the allocated intervention.

The study included intervention (NEX) and control (CON) groups with automated randomization following completion of baseline assessments. Block randomization with computer-generated random allocation sequences was employed to ensure equal distribution between intervention groups while maintaining allocation concealment. Variable block sizes of 4, 6, and 8 participants were utilized to prevent prediction of future allocations while maintaining approximate balance between groups throughout the recruitment period. The randomization sequence was generated using Robust Randomization App (RRApp) (Tu & Benn, 2017), with allocation concealment maintained until participant enrollment. Randomization was executed automatically following completion of baseline assessments and informed consent procedures, with immediate notification of group assignment to research staff.

Due to the inherently different nature of the NExGEN personalized prompt system versus the structured manual ChatGPT approach, participants could readily identify their allocated intervention, making participant blinding unfeasible. This limitation is consistent with most digital health intervention trials where the intervention modality is apparent to users. Research personnel conducting outcome assessments and data analysts remained blinded to group allocation throughout data collection and initial analysis phases to minimize assessment bias and ensure objective measurement of endpoints.

### 2.3 Participants

Adults aged 18 to 65 years residing in Greater Kuala Lumpur (Klang Valley), Malaysia, were eligible for inclusion to ensure homogeneity of the study population and facilitate standardized follow-up procedures. The inclusion criteria included a body mass index (BMI) between 27.5 and 34.9 kg/m^2^, representing individuals with overweight to moderate obesity who could safely participate in lifestyle modification interventions. Furthermore, participants were required to be in stable physical health without medical conditions that could be adversely affected by weight reduction or that might interfere with the study objectives, as determined by medical screening. Individuals with existing health conditions or concurrent medications were required to provide medical clearance from their healthcare provider confirming safety and appropriateness for participation in a weight management program. Given the digital nature of both interventions utilizing ChatGPT platforms, participants were required to demonstrate basic computer literacy and smartphone proficiency necessary for effective engagement with the AI-based systems. This included ability to navigate chat interfaces, input text responses, and maintain consistent interaction with the digital platform. Sample size calculations were based on effect sizes from comparable trials (Chambliss et al., 2011; Davies et al., 2012; Sherrington et al., 2016; Whittaker et al., 2016), with power analysis indicating that 140 participants (70 per group) would provide 80% power to detect a clinically meaningful between-group difference of 3 kg (α=0.05), accounting for an anticipated 20% dropout rate.

Participants were recruited through a multi-channel approach including social media advertisements, community outreach programs, and local news announcements to ensure diverse representation. Through the various recruitment channels, interested individuals accessed a secure online registration portal where they completed eligibility screening questionnaires, provided preliminary demographic information, and scheduled initial assessment appointments with research coordinators. Written informed consent was obtained from all participants following comprehensive explanation of study procedures, potential risks and benefits, data handling practices, and participants’ rights to withdraw at any time without penalty. During the registration process, information about the study was provided, inclusion and exclusion criteria were verified, and appointments were scheduled for baseline assessments. Final enrollment was confirmed upon successful completion of baseline assessments, medical clearance verification, and randomization assignment, with participants receiving their allocated intervention materials and initial training. To enhance engagement and provide access to premium AI capabilities, all study participants received complimentary 6-month subscriptions to ChatGPT Team accounts, which served both as an incentive for participation and as the essential technological platform required for implementing the NExGEN system and structured manual interventions.

### 2.4 Intervention

Both intervention arms were designed as fully automated digital health solutions requiring no direct human counselor involvement during the 12-week intervention period, allowing for scalable implementation and standardized delivery across participants.

The NExGEN system represents a digital platform designed to create personalized prompts for ChatGPT-based weight management interventions. This system captures interpersonal variabilities through 111 assessment questions encompassing nutrition status, fitness levels, demographics, medical history, sleep patterns, stress levels, and motivational factors. The NExGEN-ChatGPT system underwent validation using the fuzzy Delphi method to establish expert consensus on generated plan quality, with detailed findings currently under publication review and available as a pre-print on medRxiv (MEDRXIV/2025/334528). Participants complete comprehensive assessments covering goal setting, physiological measurements, dietary preferences, exercise equipment access, and environmental factors to enable highly personalized intervention strategies. Participants were then instructed to copy this combined information which consist of detailed personal attributes alongside structured instructions into ChatGPT (using o3 model; the highest reasoning model at the time of data collection), thus enabling the chatbot to function effectively as a personal trainer, nutritionist, and health advisor. The system operates through six sequential prompts that guide ChatGPT to deliver structured weight management support. These prompts establish ChatGPT as a health specialist, generate personalized behavioral recommendations, create weekly exercise plans, develop seven-day dietary plans, produce grocery shopping lists, and provide meal preparation guides. Each prompt builds upon previous responses while incorporating individual participant data to ensure culturally appropriate and locally relevant recommendations. This systematic approach enables scalable delivery of personalized nutrition and exercise guidance through automated AI-driven interactions.

The intervention group received the NExGEN system, which provided personalized prompt generation for ChatGPT based on comprehensive individual assessment data. The NExGEN system utilized an algorithm-driven approach to create individualized prompts based on 111 assessment variables including demographics, health status, dietary preferences, physical activity levels, motivational factors, and environmental considerations. To ensure research validity, the NExGEN system was standardized across all intervention participants, with identical assessment protocols and prompt generation algorithms applied consistently throughout the study period. The intervention was designed as a 12-week intensive phase followed by a 12-week maintenance period during which participants retained access to their allocated systems for continued support and guidance.

The 12-week intervention period was organized into three distinct phases to facilitate systematic behavior change and skill development, with each phase building upon previous learning and adaptation. During the initial exploration phase (weeks 1-3), participants familiarized themselves with their allocated system, learned to navigate the NExGEN and ChatGPT interface effectively, and began establishing foundational habits for sustainable lifestyle modification. The consolidation phase (weeks 4-8) focused on reinforcing newly acquired behaviors, refining personalized strategies based on early experiences, and addressing initial challenges to promote sustained engagement with recommended interventions. In the final reinforcement phase (weeks 9-12), emphasis was placed on strengthening established healthy behaviors, developing long-term maintenance strategies, and preparing participants for independent continued application of learned principles. Despite the structured phase framework, both intervention systems allowed participants to access all features and content at their preferred pace and frequency, accommodating individual schedules, learning styles, and changing needs throughout the intervention period. All system features and capabilities remained continuously available to participants, enabling self-directed learning and intervention adaptation based on personal progress, preferences, and emerging circumstances.

The core intervention strategy emphasized sustainable lifestyle modification through evidence-based approaches to dietary optimization and physical activity enhancement, rather than pursuing rapid weight loss through restrictive methods. Rather than promoting aggressive caloric restriction or rapid weight reduction approaches, the intervention prioritized gradual, sustainable weight loss strategies that emphasized long-term behavior change and metabolic health preservation. The intervention targeted moderate weight reduction of 0.5-1.0 kg per week through balanced energy deficit creation, prioritizing fat mass reduction while preserving lean body mass and supporting metabolic rate maintenance for long-term success (Ashtary-Larky et al., 2020). Participants were guided to establish individualized weight loss targets ranging from 5-10% of baseline body weight over the 12-week intervention period, with goals adjusted based on personal preferences, medical considerations, and progress trajectories. While target weight loss ranges were established, participants achieving greater weight reduction through healthy means were supported and monitored for safety, with no artificial limitations imposed on successful outcomes.

The NExGEN system incorporated personalized activity recommendations including specific exercise modalities, duration targets, intensity levels, and progression schedules tailored to individual fitness levels, preferences, and equipment availability. Typical personalized recommendations included structured walking programs, bodyweight resistance exercises, flexibility routines, recreational activities aligned with personal interests, and adaptive modifications for varying fitness levels and physical limitations. System interactivity was generated through dynamic prompt-response cycles with ChatGPT, personalized feedback based on self-reported progress, adaptive recommendation adjustments, and ongoing dialogue about challenges, successes, and strategy refinements. Participants received structured weekly objectives including specific behavioral goals, self-monitoring targets, educational modules, and skill-building activities designed to promote systematic progression toward long-term lifestyle change. The adherence level was collected through daily data logging of the dietary and physical exercise in the chat dialogue. The system provided personalized grocery shopping lists based on individual dietary plans and preferences, along with culturally appropriate recipe collections adapted for local ingredient availability and cooking methods common in Malaysia.

The CON group received structured manual prompts for ChatGPT designed to provide general weight management guidance without personalized assessment-based customization, representing standard digital health information delivery. CON participants accessed structured manual prompts covering general weight management topics including basic nutrition education, exercise recommendations, and behavioral strategies, delivered through the same ChatGPT platform for consistency. The CON group were provided static, non-adaptive content that remained consistent across all participants without individualization based on personal characteristics, preferences, or progress feedback.

Following completion of the 12-week active intervention period, participants in both groups retained access to their respective systems during the 12-week follow-up phase to support maintenance of achieved benefits and continued behavior change reinforcement. Continued system access during the follow-up period allowed for assessment of intervention durability, maintenance strategy effectiveness, and long-term user engagement patterns with the digital health tools.

### 2.5 Outcome Variables

The primary outcome of the study was body weight. Body weight was assessed using the validated Seca mBCA 515 bioelectrical impedance analysis system (Seca GmbH & Co KG), which provides accurate and reliable measurements for research applications with established validity in clinical populations. Standardized weight measurements were obtained with participants wearing minimal clothing (underwear only), having removed all accessories including jewelry, glasses, and electronic devices, following a 12-hour overnight fast, and with empty bladder to ensure measurement precision and reproducibility.

Additional outcome measures encompassed behavioral modifications (dietary intake patterns and physical activity levels) and physiological parameters (body composition, cardiometabolic markers, and cardiovascular health indicators) to provide comprehensive assessment of intervention effects. Seven-day dietary records were generated at each of the 4 measurement time points using Nutritionist Pro™ software (version 7.9, Axxya Systems, Stafford, TX, USA), with reference to the Malaysia Food Composition Table for local food items (Tay et al., 2023). Comprehensive nutritional analysis included assessment of total energy intake, macronutrient distribution (carbohydrates, proteins, fats), dietary energy density excluding beverages, micronutrient adequacy, and food group consumption patterns to evaluate intervention-induced dietary changes. Objective physical activity measurement was conducted using validated ActiGraph GT3X+ accelerometers (ActiGraph LLC, Pensacola, USA) worn at the waist for seven consecutive days, supplemented by the International Physical Activity Questionnaire-Long Form (IPAQ-LV) to capture subjective activity perceptions and domains not captured by accelerometry (Ahmad et al., 2018; Wanner et al., 2016). A minimum threshold of five valid days (including at least one weekend day) with ≥10 hours of wear time was required for accelerometer data inclusion, while dietary records required completion of at least five days (including one weekend day) to ensure representative assessment of habitual patterns (Chomistek et al., 2017).

Comprehensive body composition assessment included measurement of body height using a calibrated stadiometer (Seca 274), waist circumference using standardized techniques with a non-stretchable measuring tape (Seca 201), and detailed body composition analysis (fat mass, fat-free mass, body fat percentage) via bioelectrical impedance analysis using the Seca mBCA 515 system (Chamberlin et al., 2025; Stratton et al., 2021; Wu et al., 2024). Blood samples were collected for analysis of glycated hemoglobin (HbA1c) and blood lipids (total cholesterol, high-density lipoprotein cholesterol, low-density lipoprotein cholesterol, and triglycerides) using the Cobas b 101 system (Roche Diagnostics GmbH, Mannheim, Germany) (Lenters-Westra & English, 2021; Li et al., 2023). Fasting glucose samples were analyzed using the HemoCue HB 201 System (HemoCue, Angelholm, Sweden) (Méndez-Gómez-Humarán et al., 2024). Blood pressure was measured using a validated validated oscillometric device (Dinamap 420, GE Healthcare, Chicago, IL) according to standardized protocols (Carey & Whelton, 2018; Jones et al., 2025).

### 2.6 Data Analysis

IBM SPSS Statistics (Version 30) and Graphpad Prism (Version 10.5) were used for statistical analysis and creation of graphs. Statistical analysis was conducted using linear mixed models for repeated measures (MMRM) to account for the longitudinal study design, within-subject correlations, and missing data patterns, with treatment group, time, and group-by-time interactions as fixed effects. The significance level was set at .05 for all comparisons. Graphs of descriptive results of anthropometric variables were created using Graphpad Prism (Version 10.5).

Per-protocol (PP) and intention-to-treat (ITT) analyses were performed (Tripepi et al., 2020). The per-protocol analysis included only participants who completed the study without major protocol violations and had available outcome data for the specific variable being analyzed, providing insight into intervention efficacy under optimal adherence conditions (Molero-Calafell et al., 2024). When outcome data were missing for specific variables, the corresponding participants were excluded from those particular analyses, resulting in variable sample sizes across different outcome measures depending on data completeness. The intention-to-treat analysis incorporated all randomized participants according to their original group assignment, regardless of actual intervention adherence or study completion status, providing an assessment of intervention effectiveness under real-world conditions (Tripepi et al., 2020; Molero-Calafell et al., 2024). Missing data in the ITT analysis were addressed using multiple imputation techniques (*m*=50 imputations) under the missing-at-random assumption (Austin et al., 2021; Jakobsen et al., 2017), with imputation models including baseline characteristics, treatment group, and available follow-up measurements.

Per-protocol and intention-to-treat analyses yielded comparable results across all measured variables, suggesting robust intervention effects that were not substantially influenced by missing data or protocol adherence variations (Hernán & Robins, 2017). Given the comprehensive nature of the outcome variables assessed and to provide the most conservative effectiveness estimates, results from the intention-to-treat analysis are primarily presented, representing the pragmatic impact of offering these interventions to eligible populations (Tripepi et al., 2020).

### 2.7 Ethical Considerations

This study followed the principles of the Declaration of Helsinki. The study was approved by the Universiti Teknologi MARA Institutional Review Board (Ethics REC/09/2024 (PG/FB/30)). A preliminary feasibility study was conducted with a subset of participants to evaluate recruitment procedures, intervention delivery, and outcome measurement protocols, with no modifications required to the main study protocol based on pilot findings. Comprehensive written informed consent was obtained from all participants prior to enrollment, following detailed explanation of study procedures, potential risks and benefits, data confidentiality measures, and participants’ rights to withdraw from the study at any time without affecting their future healthcare or relationships with the research institution.

## 3. Results

### 3.1 Participants

Recruitment of participants who were living in Greater Kuala Lumpur (Klang Valley), from July 2024 to December 2024, took place online and offline through various media, such as news items, flyers, and social media. During this extended recruitment period, a total of 257 potential participants formally expressed their interest in the study. After screening (telephone interview and preliminary examination), 160 subjects were included in the study, and participants were sequentially booked for baseline assessments. Throughout the study duration, there were 33 dropouts. None of the health reasons of the dropouts were associated with the intervention, indicating that participant withdrawal was not attributable to adverse effects or intervention-related complications. The study participant flow is shown in Figure 1. The baseline characteristics of the included participants are summarized in Table 1.

**Figure 1:**
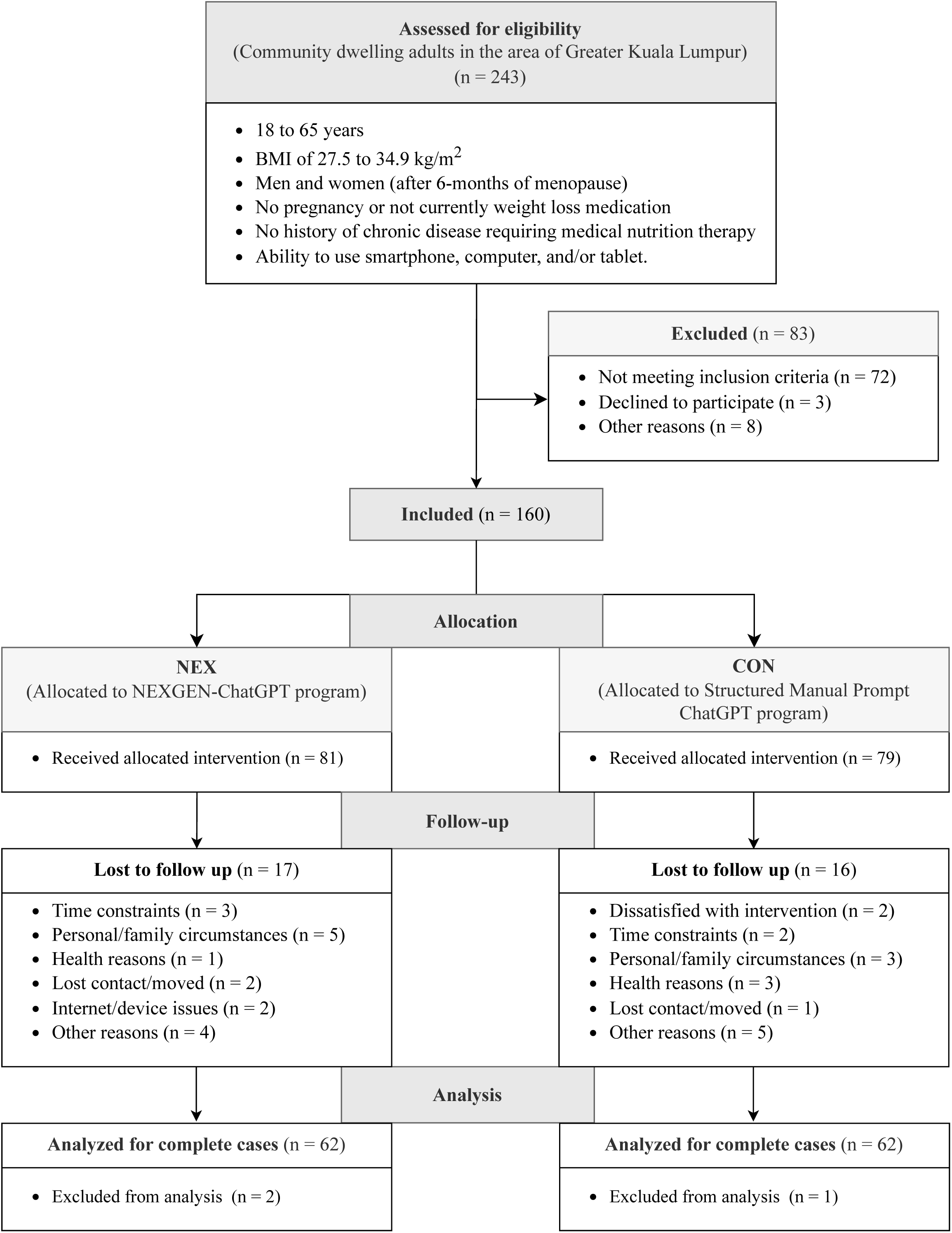
CONSORT Flow chart depicting participant recruitment, allocation, and dropout during 12-week intervention and 12-week follow-up.

### 3.2 Weight and Body Composition Changes

Body weight (Fig. 2A) was significantly reduced by the NEX intervention as indicated by the response across time differing between groups (P < 0.001; effect size, 0.14). Follow-up tests show body weight was significantly lowered in NEX from baseline at 12-week and follow-up (P < 0.001 for both), representing reductions of 6.6 kg (7.5%) and 5.5 kg (6.2%), respectively. Between groups, NEX achieved significantly greater weight loss than CON at 12-week (mean difference −3.2 kg; P = 0.016) and follow-up (mean difference −3.5 kg; P = 0.018). Waist circumference (Fig. 2B) showed a significant time × intervention interaction (P = 0.000; effect size, 0.13), with NEX demonstrating reductions of 5.4 cm (5.3%) at 12-week and 5.0 cm (4.9%) at follow-up from baseline (both P < 0.001).

**Figure 2:**
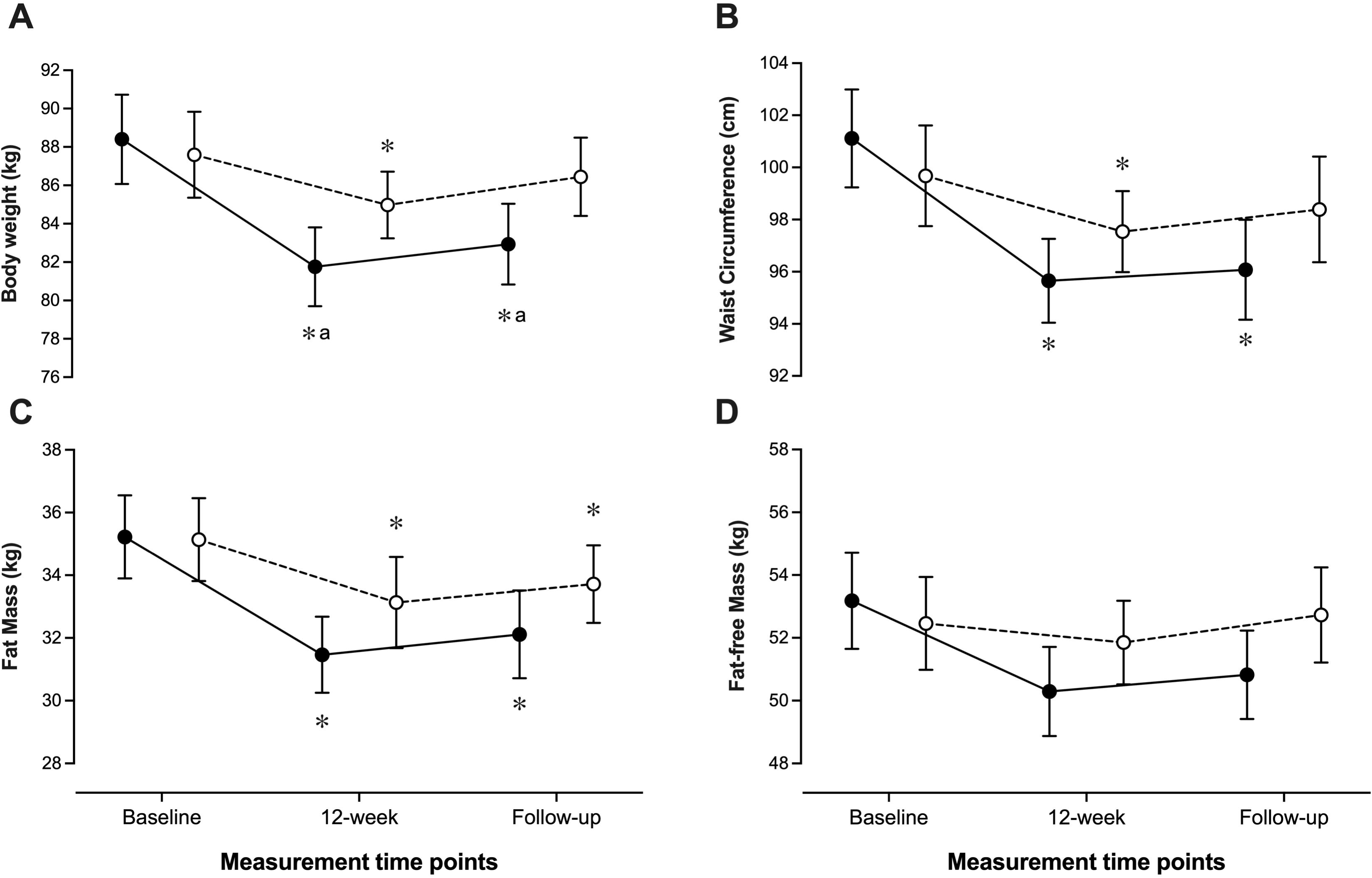
Data (mean and 95% CI) of body composition variables in the NEX (●) and CON (○) groups (intention-to-treat analysis). The time points were baseline, after the 12-week intervention, and after an additional 3 months of follow-up. (A) Body weight; (B) Fat mass; (C) Waist circumference; (D) Fat-free mass. *Difference from baseline (P < 0.05); ^a^difference from CON (P < 0.05).

Fat mass (Fig. 2C) was significantly reduced by the NEX intervention (P = 0.02; effect size, 0.10), with decreases of 3.7 kg (10.5%) at 12-week and 3.1 kg (8.8%) at follow-up compared to baseline (both P < 0.001). Between-group differences showed NEX achieved 1.7 kg greater fat loss than CON at 12-week (P = 0.082) and 1.6 kg greater loss at follow-up (P = 0.097). Fat-free mass (Fig. 2D) remained stable across all time points with no significant main effects or interactions (P > 0.05 for all comparisons), indicating that weight loss was primarily from adipose tissue. The CON group experienced gradual increases in body weight (3.0% at 12-week, 1.7% at follow-up), fat mass (5.9% at 12-week, 4.0% at follow-up), and waist circumference (2.1% at 12-week) before returning near baseline at follow-up.

### 3.3 Dietary and Physical Activity Changes

At baseline prior to any intervention, the NEX group had a mean energy density of 4.84 ± 0.25 kcal/g, energy intake of 1833.2 ± 659.7 kcal/day, and fat intake of 80.9 ± 33.0 g/day, while the CON group showed comparable values of 4.84 ± 0.26 kcal/g, 1830.1 ± 656.7 kcal/day, and 80.5 ± 31.9 g/day, respectively. Energy density (Fig. 3A) was significantly modified by NEX intervention as indicated by the response across time differing between groups (P = 0.0014; effect size, 0.041). Follow-up tests showed energy density was significantly lower in NEX compared to baseline at 12-week (P < 0.001) and follow-up (P = 0.0005). Compared with CON, NEX showed significantly lower energy density at 12-week (95% CI −0.213 to −0.076; P < 0.001) and follow-up (95% CI −0.203 to −0.008; P = 0.0005). Fat intake (Fig. 3E) was reduced by NEX intervention as indicated by the response across time differing between groups (P < 0.001; effect size, 0.066). Follow-up tests showed fat intake was significantly reduced in NEX from baseline at 12-week (95% CI 9.53 to 20.83; P < 0.001) and follow-up (95% CI 5.57 to 18.07; P < 0.001). Compared with CON, NEX was significantly lower at 12-week (95% CI −16.85 to −0.152; P = 0.046) and follow-up (95% CI −19.30 to −2.10; P = 0.015). No significant changes in these parameters were observed during CON intervention.

**Figure 3:**
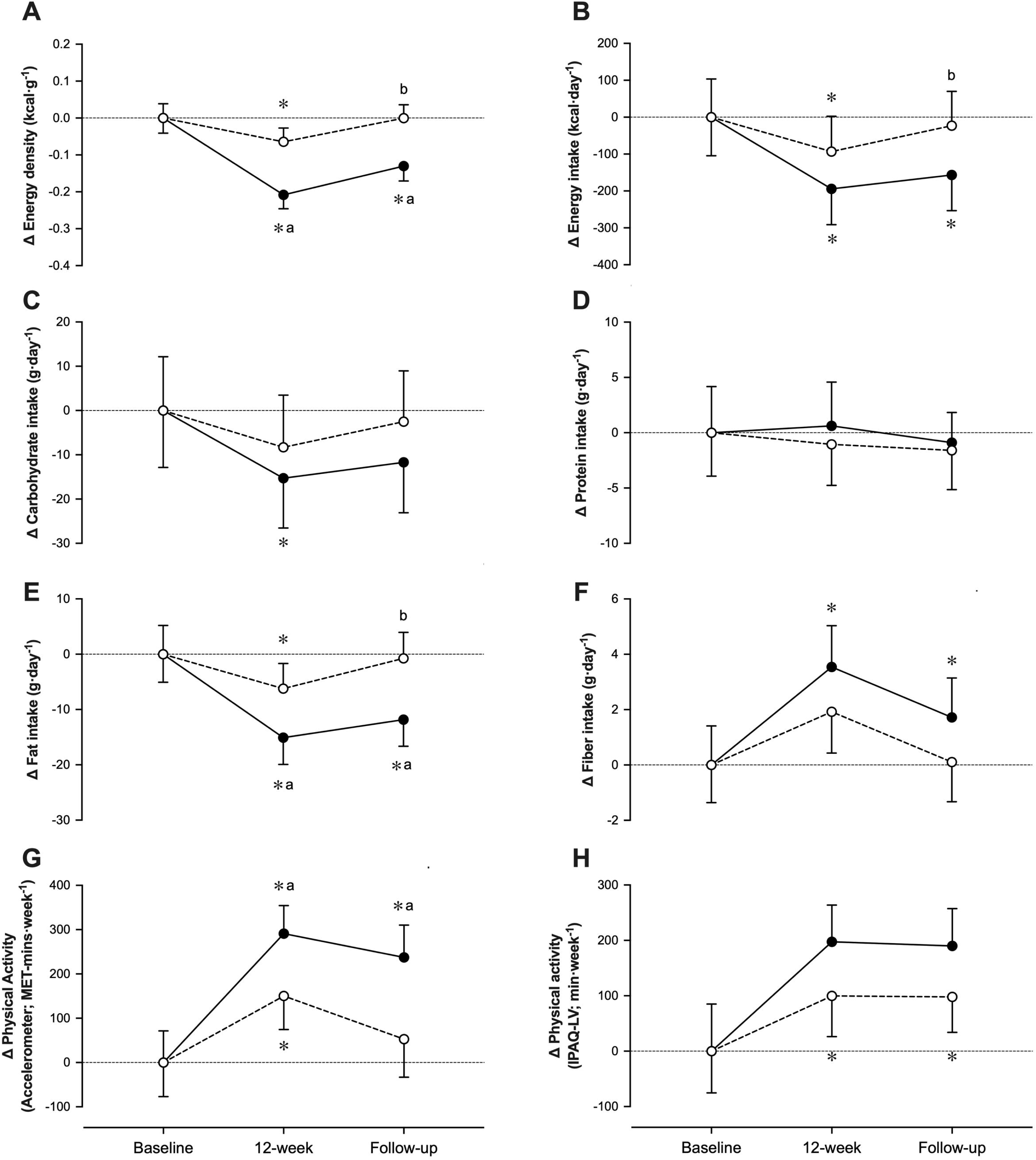
Change (Δ) relative to baseline in energy density (A), energy intake (B), carbohydrate intake (C), protein intake (D), fat intake (E), fiber intake (F), physical activity measured by accelerometer (G), and physical activity measured by IPAQ-LV (H), during the 12-week intervention period and 3-month follow-up with NEX (●) and CON (○) (group means ± SE) (intention-to-treat analysis). *Difference from baseline (P < 0.05); ^a^difference from CON (P < 0.05); ^b^difference from 12-week (P < 0.05).

Fiber intake (Fig. 3F) increased significantly across time points (P < 0.001; effect size, 0.128), with both groups showing improvements from baseline. The NEX group demonstrated significant increases from baseline at 12-week (95% CI −5.70 to 4.46; P < 0.001) and follow-up (95% CI −4.27 to 6.06; P < 0.001). The CON group also showed significant improvements, though the time × intervention interaction was not statistically significant (P = 0.076). Energy intake and carbohydrate intake showed significant time effects (P < 0.001 and P = 0.0117, respectively) with reductions observed in both groups, but no significant between-group differences were detected. Protein intake remained stable across all time points with no significant main effects or interactions observed (P > 0.05 for all comparisons). The mean difference between groups for energy intake was −75.16 kcal/day (95% CI −252.1 to 101.8; P = 0.403), and for carbohydrate intake was −4.863 g/day (95% CI −24.89 to 15.17; P = 0.6322).

Physical activity measured by accelerometry (Fig. 3G) was significantly enhanced by NEX intervention as indicated by the response across time differing between groups (P = 0.0017; effect size, 0.040). Follow-up tests showed accelerometer-measured physical activity was significantly increased in NEX from baseline at 12-week (95% CI −367.4 to −214.9; P < 0.001) and follow-up (95% CI −312.0 to −162.8; P < 0.001). Compared with CON, NEX was significantly higher at 12-week (95% CI 20.6 to 277.9; P = 0.023) and follow-up (95% CI 25.63 to 360.7; P = 0.0241). Similarly, self-reported physical activity using IPAQ-LV (Fig. 3H) was enhanced by NEX intervention with the response across time differing between groups (P = 0.0021; effect size, 0.038). Follow-up tests revealed significant increases in NEX from baseline at 12-week (95% CI −276.7 to −118.4; P < 0.001) and follow-up (95% CI −264.1 to −116.1; P < 0.001). Both accelerometer and self-reported measures demonstrated sustained improvements at follow-up, indicating the durability of behavioral changes induced by the NEX intervention. The CON group showed modest increases in physical activity over time, but these changes were significantly smaller than those observed in the NEX group.

### 3.4 Metabolic and Blood Pressure Changes

The cardiometabolic responses to the NEX intervention across baseline, 12-week, and 3-month follow-up assessments are summarized in Table 2. HbA1c levels remained stable throughout the study period, with no significant intervention effect observed (P = 0.82). Follow-up tests revealed no significant differences between NEX and CON groups at any time point (baseline: 95% CI −0.12 to 0.127, P = 0.949; 12-week: 95% CI −0.153 to 0.089, P = 0.607; follow-up: 95% CI −0.115 to 0.099, P = 0.879). Similarly, neither group showed significant within-group changes from baseline values across the intervention and follow-up periods.

Total cholesterol demonstrated a significant time effect (P < 0.0001; Geisser-Greenhouse’s epsilon = 0.99), with both groups showing reductions over time. In the NEX group, total cholesterol was significantly reduced from baseline at 12-week (95% CI −21.48 to 7.419, P < 0.0001) and follow-up (95% CI −20.44 to 6.813, P < 0.0001). The CON group showed a significant reduction only at follow-up (95% CI −17.94 to 0.380, P = 0.039). However, no significant between-group differences were observed at any time point. LDL cholesterol exhibited similar patterns with a significant time effect (P = 0.001), showing reductions in the NEX group at 12-week (95% CI −15.85 to 2.67, P = 0.0035) and follow-up (95% CI −14.52 to 0.856, P = 0.023), while CON group changes remained non-significant. HDL cholesterol and triglycerides showed no significant intervention effects (P = 0.014 and P = 0.548, respectively), with minimal between-group differences observed throughout the study period.

Blood pressure parameters remained largely unchanged across both groups throughout the intervention period. Systolic blood pressure showed no significant intervention effect (P = 0.402), with between-group differences remaining non-significant at all time points (baseline: 95% CI −4.317 to 3.19, P = 0.767; 12-week: 95% CI −6.24 to 1.20, P = 0.183; follow-up: 95% CI −5.27 to 2.60, P = 0.503). Within-group analyses revealed no significant changes from baseline in either group across the study duration. Diastolic blood pressure demonstrated similar stability (intervention effect P = 0.839), with consistently non-significant between-group comparisons and minimal within-group variations. Mean arterial pressure showed a significant time effect (P = 0.046), primarily driven by a small but significant reduction in the NEX group from baseline to 12-week (95% CI −2.56 to 0.141, P = 0.025). However, this change was not sustained at follow-up, and no significant between-group differences emerged at any assessment point.

## 4. Discussion

The principal finding of this randomized controlled trial was that the NExGEN personalized prompt generator integrated with ChatGPT demonstrated superior efficacy for weight reduction and body composition improvement compared to structured manual ChatGPT-based guidance in adults with overweight and obesity. The personalized AI-driven approach produced significantly greater weight loss than the CON condition, with effects sustained throughout the 24-week study period including a 12-week maintenance phase. The NExGEN system achieved 6.6 kg (7.5%) weight loss at 12 weeks, which compares favorably to reported outcomes from digital interventions. At the 24-week assessment, participants in the NExGEN group maintained 5.5 kg (6.2%) weight reduction from baseline, demonstrating sustained effectiveness beyond the active intervention period. A recent systematic review of systematic reviews found that eHealth interventions achieved statistically significant weight loss outcomes with mean differences ranging from −1.07 to −3.10 kg compared to CON conditions (Kupila et al., 2023). Digital interventions with self-monitoring components have shown promising results, with one meta-analysis demonstrating a mean difference of −2.87 kg for digital self-monitoring interventions (Berry et al., 2021). Many digital interventions face challenges with long-term maintenance due to high attrition rates and declining engagement over time (Neve et al., 2010). Digital interventions incorporating human coaching elements have achieved enhanced effectiveness over purely automated systems, though such approaches present scalability challenges and increased implementation costs (Kupila et al., 2023).

Fat mass in the NEX intervention group decreased significantly compared with the CON group over the study course, whereas fat-free mass remained stable in both groups without significant between-group differences. These findings are important because few AI-based interventions perform comprehensive body composition measurements using validated bioelectrical impedance analysis. Recent trials indicate that personalized AI-driven weight loss interventions can produce greater improvements in body composition compared to standard digital programs (Pokushalov et al., 2025). An AI-guided program leveraging individualized data led to significantly more fat mass reduction than physician-guided approaches, with participants achieving approximately 12% weight loss versus 7% in controls (Moravcová et al., 2022; Pokushalov et al., 2025). Preservation of fat-free mass during weight loss is critical because low fat-free mass is associated with higher mortality and plays an important role in energy expenditure (Moravcová et al., 2022). The NEX intervention achieved above-average reduction in waist circumference, indicating significant visceral fat loss. Achieving sustained 5-7% weight loss appears sufficient to induce meaningful improvements in fat distribution, with clinical evidence showing approximately 5% weight loss can reduce visceral adipose tissue by 10% or more (Ryan & Yockey, 2017). The selective fat loss while maintaining lean mass represents a favorable scenario for metabolic health, as visceral fat is the depot most linked to cardiometabolic risk (Moravcová et al., 2022).

The effectiveness of the NEX intervention was evident in behavioral variables, particularly energy density reduction, which emerged as a primary mechanism of weight loss. Energy density decreased significantly in the NEX group both short-term and long-term, consistent with prior research showing that lowering dietary energy density facilitates weight management (Kohl et al., 2023). Interactive, personalized programs led to significantly greater decreases in dietary energy density than non-personalized CON, with tailored feedback enhancing adherence to dietary strategies beyond what standardized digital content achieves (Kohl et al., 2023). The NEX group demonstrated significantly lower fat intake and higher fiber intake compared to baseline and CON, reflecting successful dietary quality improvements. Energy intake developed consistently with energy density changes, although effects were more pronounced at 12 weeks. The durability of dietary changes is noteworthy, as reductions in energy density were maintained at follow-up, highlighting that personalized digital interventions can instill lasting dietary habits through reinforced behavioral prompts (Kohl et al., 2023). Physical activity increased significantly in the NEX group as measured by both accelerometry and self-report. Discrepancies between objective and subjective measures reflect common issues in digital health trials, where individuals often overestimate activity due to social desirability bias (Loughnane et al., 2025). The sustained improvements in both dietary and activity behaviors indicate successful modification of key weight management mechanisms through personalized AI-driven prompts.

Despite positive effects on body weight and composition, improvements in cardiometabolic variables were modest and not significantly superior to CON group findings. This pattern likely reflects the characteristics of the study population, with many participants being metabolically healthy overweight adults at baseline, having relatively normal blood pressure, lipid, and glucose levels despite excess weight. In metabolically healthy individuals, weight loss interventions often show blunted improvements in cardiometabolic markers because there is less abnormality to correct (Blüher, 2020). Clinical measures such as LDL cholesterol, total cholesterol, and fasting glucose did not change dramatically over six months, especially in participants whose baseline values were within normal ranges (Moravcová et al., 2024). Blood pressure followed similar patterns, with participants having normal baseline values showing only minimal decreases. Small blood pressure reductions are possible with modest weight loss, but without hypertension at baseline, these changes are not always statistically significant (Ryan & Yockey, 2017). The magnitude of weight loss needed to substantially lower blood pressure or LDL cholesterol is greater when starting from normal baseline values. Triglycerides showed more favorable responses, with the NEX group demonstrating meaningful decreases consistent with weight loss-induced metabolic benefits. A 5% weight loss is considered necessary to achieve clinically relevant cardiometabolic effects (Ryan & Yockey, 2017). Less than half of NEX participants achieved more than 5% weight loss, which may explain the limited cardiometabolic improvements. The time course of changes may also be relevant, as cardiometabolic benefits often accrue gradually and might become significant with longer follow-up or larger samples (Pokushalov et al., 2025).

The findings support the theoretical rationale for personalized AI-driven interventions in weight management, affirming that tailoring content to individuals enhances outcomes. The NEX system generated individualized exercise and dietary prompts by integrating comprehensive assessment data, grounded in hypotheses that interventions aligned to personal context are more motivating and sustainable (Michie et al., 2017). The superior weight loss and behavioral changes achieved by the NEX group compared to standardized controls support research showing tailored interventions lead to better outcomes than non-personalized approaches (Kohl et al., 2023). Modern AI can produce high-quality, empathetic coaching text that users find acceptable, with AI-generated messages rated similarly helpful as human coach messages (Huang et al., 2025).

Several limitations must be considered in this study. First, complete blinding was not feasible due to the inherently different nature of the NExGEN personalized prompt system versus the structured manual ChatGPT approach, allowing participants to readily identify their allocated intervention modality. Therefore, the motivation and engagement of participants may have been differentially influenced by their awareness of receiving either the personalized system or standardized prompts, potentially creating expectation bias. Second, both study groups may have been additionally motivated to achieve their health goals through interest in the complimentary 6-month ChatGPT Team subscriptions provided as participation incentives, though this technological access was essential for implementing both intervention modalities. Third, suboptimal utilization of the allocated intervention systems may have attenuated the true effectiveness of the programs and influenced study results. However, usage analysis demonstrated that the NExGEN system was actively utilized by participants throughout the study period, with higher engagement and adherence levels compared to the CON group (data not shown). Fourth, because the CON group received only structured manual prompts, minimal use was expected compared to the personalized system. Despite these limitations, this study provides robust evidence that personalized AI-driven prompt generation produces significantly greater improvements in weight loss and behavioral outcomes. The rigorous randomized controlled design with standardized clinical assessments, objective physical activity measurement, and extended follow-up period represent significant methodological strengths. Given the scalability of automated AI-driven interventions, the demonstrated effectiveness may offer substantial public health benefits for addressing obesity through cost-effective digital health solutions deployable at population scale.

## 5. Conclusion

This study demonstrated that a personalized AI-driven prompt generator integrated with ChatGPT was effective and superior to structured manual ChatGPT guidance for weight management in adults with overweight and obesity. The personalization of AI-based interventions appears to play a crucial role in enhancing effectiveness beyond standardized digital approaches. Based on the present findings, generic automated prompts are insufficient to induce optimal weight loss and sustained behavioral modifications. In the personalized intervention studied here, effectiveness was achieved through individualized assessment-driven prompt generation that tailored recommendations to personal characteristics, preferences, and environmental factors. Future research should investigate which personalization elements are most critical for maximizing AI-driven weight loss intervention effectiveness. Nevertheless, the results indicate that integrating comprehensive personal assessment data into automated AI prompt generation systems represents a promising approach for delivering scalable, effective weight management interventions.

## Declarations

### Ethical approval

The study was approved by the Universiti Teknologi MARA Institutional Review Board (Ethics REC/09/2024 (PG/FB/30)).

### Informed consent

Comprehensive written informed consent was obtained from all participants prior to enrollment, following detailed explanation of study procedures, potential risks and benefits, data confidentiality measures, and participants’ rights to withdraw from the study at any time without affecting their future healthcare or relationships with the research institution.

### Author Contributions

Azwa Suraya Mohd Dan and Adam Linoby: Conceived and designed the trials; Performed the trials; Analyzed and interpreted the data; Contributed to data analysis and interpretation of the data; In charge of the project administration; Wrote the first draft of the manuscript. Sazzli Shahlan Kasim, Sufyan Zaki, and Razif Sazali: Conceived and designed the study; Interpreted the data. Yusandra Md Yusoff, Zulqarnain Nasir, and Amrun Haziq Abidin: Performed the trials; Contributed to data analyses.

### Data Availability Statement

Data will be made available from the corresponding author on reasonable request.

### Clinical Trial Number

not applicable.

## Tables

**Table 1:** Baseline demographic and clinical characteristics of study participants.

**Table 2:** Metabolic and cardiovascular parameters measured at baseline and across 12 weeks of NEX intervention and 3-month follow-up in intervention (NEX) and control (CON) groups (intention-to-treat analysis).

