## Supplementary material for "Personalized AI Prompt Generator and ChatGPT for Weight Loss: Randomized Controlled Trial in Adults with Overweight and Obesity": Table 1

**Table 1. Baseline demographic and clinical characteristics of study participants**

| **Variable** | **All participants** (N=160) | **NEX** (n=81) | **CON** (n=79) |
| --- | --- | --- | --- |
| **Age (yrs)^a^** | 35.90 ± 9.39 | 36.04 ± 9.48 | 35.75 ± 9.34 |
| **Sex^b^** |  |  |  |
| Male | 52 (32.5%) | 25 (30.9%) | 27 (34.2%) |
| Female | 108 (67.5%) | 56 (69.1%) | 52 (65.8%) |
| **Body weight (kg)^a^** | 88.01 ± 10.24 | 88.41 ± 10.54 | 87.60 ± 9.97 |
| **Body height (m)^a^** | 1.66 ± 0.10 | 1.66 ± 0.10 | 1.65 ± 0.10 |
| **BMI (kg/m^2^)^a^** | 31.20 ± 2.58 | 31.28 ± 2.60 | 31.12 ± 2.57 |
| **Dropouts^b^** | 33 (20.63%) | 17 (20.99%) | 16 (20.25%) |

^a^ values are presented as the mean ± SD. ^b^ values are presented as the n (%).
