## Supplementary material for "Personalized AI Prompt Generator and ChatGPT for Weight Loss: Randomized Controlled Trial in Adults with Overweight and Obesity": Table 2

**Table 2. Metabolic and cardiovascular parameters measured at baseline and across 12 weeks of NEX intervention and 3-month follow-up in intervention (NEX) and control (CON) groups (intention-to-treat analysis).**

| **Variable and group** | **Baseline**  **(mean ± SD)** | **12 Week**  **(mean ± SD)** | **Follow up**  **(mean ± SD)** |
| --- | --- | --- | --- |
| **HbA1c (%)** |  |  |  |
| NEX | 5.36 ± 0.35 | 5.30 ± 0.37 | 5.33 ± 0.31 |
| CON | 5.35 ± 0.43 | 5.34 ± 0.41 | 5.34 ± 0.37 |
| **Total Cholesterol (mg/dL)** |  |  |  |
| NEX | 212.6 ± 38.0 | 198.1 ± 40.7* | 199.0 ± 36.4* |
| CON | 210.3 ± 42.4 | 201.4 ± 42.0 | 201.1 ± 43.3* |
| **LDL Cholesterol (mg/dL)** |  |  |  |
| NEX | 133.2 ± 34.6 | 123.9 ± 32.5* | 125.5 ± 32.1* |
| CON | 137.7 ± 37.3 | 132.1 ± 37.1 | 132.4 ± 39.9 |
| **HDL Cholesterol (mg/dL)** |  |  |  |
| NEX | 56.1 ± 25.7 | 52.9 ± 24.5 | 51.5 ± 25.7 |
| CON | 48.8 ± 30.8 | 46.4 ± 28.7 | 45.9 ± 33.6 |
| **Triglycerides (mg/dL)** |  |  |  |
| NEX | 116.6 ± 56.3 | 106.6 ± 48.0* | 109.7 ± 56.6 |
| CON | 118.7 ± 61.9 | 114.4 ± 48.6 | 114.0 ± 50.8 |
| **Systolic blood pressure (mmHg)** |  |  |  |
| NEX | 127.9 ± 11.7 | 126.2 ± 11.5 | 128.4 ± 11.6 |
| CON | 128.5 ± 12.3 | 128.7 ± 12.3 | 129.7 ± 13.5 |
| **Diastolic blood pressure (mmHg)** |  |  |  |
| NEX | 88.4 ± 9.2 | 87.2 ± 8.4 | 87.0 ± 8.5 |
| CON | 87.9 ± 9.0 | 87.2 ± 8.8 | 88.2 ± 8.5 |
| **Mean Arterial pressure (mmHg)** |  |  |  |
| NEX | 101.5 ± 9.4 | 100.2 ± 8.8* | 100.8 ± 8.9 |
| CON | 101.4 ± 9.6 | 101.1 ± 9.3 | 102.0 ± 9.5 |

Values are means ± SD. *Different from baseline, *P* < 0.05
